# Coronavirus (COVID-19) Outbreak Prediction Using Epidemiological Models of Richards Gompertz Logistic Ratkowsky and SIRD

**DOI:** 10.1101/2020.11.29.20240580

**Authors:** Ahmad Sedaghat, Seyed Amir Abbas Oloomi, Mahdi Ashtian Malayer, Nima Rezaei, Amir Mosavi

## Abstract

On 30 July 2020, a total number of 301,530 diagnosed COVID-19 cases were reported in Iran, with 261,200 recovered and 16,569 dead. The COVID-19 pandemic started with 2 patients in Qom city in Iran on 20 February 2020. Accurate prediction of the end of the COVID-19 pandemic and the total number of populations affected is challenging. In this study, several widely used models, including Richards, Gompertz, Logistic, Ratkowsky, and SIRD models, are used to project dynamics of the COVID-19 pandemic in the future of Iran by fitting the present and the past clinical data. Iran is the only country facing a second wave of COVID-19 infections, which makes its data difficult to analyze. The present study’s main contribution is to forecast the near-future of COVID-19 trends to allow non-pharmacological interventions (NPI) by public health authorities and/or government policymakers. We have divided the COVID-19 pandemic in Iran into two waves, Wave I, from February 20, 2020 to May 4, 2020, and Wave II from May 5, 2020, to the present. Two statistical methods, i.e., Pearson correlation coefficient (R) and the coefficient of determination (R2), are used to assess the accuracy of studied models. Results for Wave I Logistic, Ratkowsky, and SIRD models have correctly fitted COVID-19 data in Iran. SIRD model has fitted the first peak of infection very closely on April 6, 2020, with 34,447 cases (The actual peak day was April 7, 2020, with 30,387 active infected patients) with the re-production number R0=3.95. Results of Wave II indicate that the SIRD model has precisely fitted with the second peak of infection, which was on June 20, 2020, with 19,088 active infected cases compared with the actual peak day on June 21, 2020, with 17,644 cases. In Wave II, the re-production number R0=1.45 is reduced, indicating a lower transmission rate. We aimed to provide even a rough project future trends of COVID-19 in Iran for NPI decisions. Between 180,000 to 250,000 infected cases and a death toll of between 6,000 to 65,000 cases are expected in Wave II of COVID-19 in Iran. There is currently no analytical method to project more waves of COVID-19 beyond Wave II.

## I. Introductions

Severe acute respiratory syndrome coronavirus 2 (SARS-CoV-2) which is responsible for novel coronavirus disease (COVID-19) was first observed in Wuhan-China in late December 2019 [1,2]. By 4 June 2020, COVID-19 was spread worldwide in 215 countries with 6,638,912 infected cases, 3,204,292 recovered, and 389,816 deceased cases [3]. The outbreak of COVID-19 in Iran was first observed in the City of Qom, a pilgrims’ center in Iran, which promptly extended to Tehran. Tehran, the Capital of Iran, is currently the center of outbreaks [4]. The study by Ahmadi et al. [4] suggested the dense population in Tehran and nearby cities stemmed from high initial growth rates of 36.3 to 161.6 in Iran. The Iranian government then implemented measures on banning movement between densely populated provinces and cities as an effective method to reduce the spread of the COVID-19 outbreaks. The maximum infection day was 3,186 cases on 30 March 2020. On 31 May 2020, 151,491 individuals were reported to be infected. Among them, 118,848 recovered, and 7,804 died in Iran [3].

Lin et al. [2] applied several predictive models, including Gompertz, Logistic, Bertalanffy, and Gompertz, to study COVID-19 in China. Roda et al. [5] explained why it is hard to predict a pandemic from a mathematical point of view. Zhang et al. [6] studied the duration and outbreak of COVID-19 in several countries in Europe and America, using power-law and exponential law models. Li et al. [7] applied SIR mixture model to tackle double peak epidemy situations. The method is cumbersome and not easy to implement. Mummert et al. [8] proposed a simpler multiple-waves model to study pandemics. However, it is not clear if the method can be implemented in typical ODE equation models, such as the SIR method. Another possible candidate for a double peak pandemic may be found in Sazonov et al. [9], yet, the method is cumbersome and complex.

In the present study, we have analyzed several widely used methods in epidemiology or microbiology, such as Richards, Gompertz, Logistic, and Ratkowsky, to fit present and previous trends of COVID-19 in Iran. We applied the Pearson correlation coefficient (R) and the coefficient of determination (R2) with the aid of an optimization tool in MATLAB to evaluate the goodness of fitted data. Results of the studied models are used to project future trends of COVID-19 in Iran, including the total number and the end date of different COVID-19 populations, i.e., infected, recovered, and dead population. The objective is not to meddle with these models analytically, instead to get the best of these models using optimization to find their optimized parameters and to provide further insight to health authorities or policymakers on non-pharmacological interventions (NPI) decisions.

## II. Materials and methdos

*SIR* The COVID-19 pandemic outbreak in Iran started on 20 February 2020 with 2 infected cases in Qom city. Qom city has approximately 1.2 million populations and is considered as wholly land for Shia Muslims with several pilgrimage shrines in Iran. The city’s social structure and travelers into and from the city have made COVID-19 propagation in Iran unique and hard to analyze. In the present study, we have applied several mathematical models widely used in epidemiological studies, including Richards, Gompertz, Logistic, Ratkowsky, and SIRD models, to understand dynamics of COVID-19 in Iran better. The Richards, Gompertz, Logistic, and Ratkowsky models only deal with single population; therefore, we have used each model three times to obtain fitting functions for infected, recovered, and deceased clinical data. Obtaining parameters of each model is not easy task and has to be done using programming in MATLAB; although the results of fitting parameters are initial value dependent and should be obtained carefully.

### A. General Epidemiology models

We have adopted several widely used models, which use exponential functions to express the nature of endemic/pandemic fast-growing contagious diseases. Follow are listed these models [10-13]. We have divided COVID-19 clinical data into two waves, because only one peak can be dealt by the models described here. Wave I finish after passing first peak and reaching to the minimum value. Wave 2 starts from the minimum value of Wave I. To obtain the parameters of these models, we adopted an optimization technique to minimize error between the model values and clinical data. In literature, some constant numbers were suggested for these models which are not necessarily the best fit coefficients.

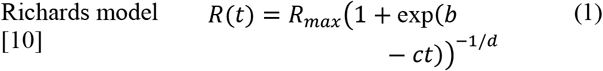

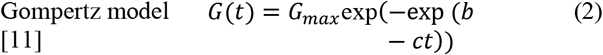

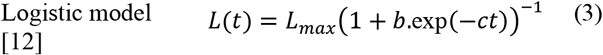

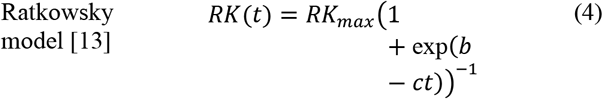

In equation (1), *R*(*t*) expresses temporal change of a pandemic population such as infected, recovered, deceased and so on. *R*_*max*_ is the maximum size of studied population, parameters b, c, and d are model parameters and time, t, is usually expressed by number of days from outbreak of infectious disease.

In equation (2), *G*(*t*) is temporal function of a pandemic population such as infected, recovered, deceased and so on. *G*_*max*_ is the maximum size of population and parameters, b and c, are model constants.

In equation (3), *L*(*t*) is Logistic function of time (t), *L*_*max*_ is the maximum limit of pandemic population, and b and c are model parameters.

In equation (4), *RK*(*t*) is the Ratkowsky function of time. *RK*_*max*_ is the maximum value of the pandemic population. Ratkowsky constants b and c are found by fitting with data.

### B. SIRD Model

SIRD model is an extension of SIR model first proposed by Kermack and McKendrick [14] to study outbreak of contagious epidemics such as plague and cholera [6]. In the SIR model, it is assumed that the population’s size is fixed and remain constant throughout epidemy. It is also assumed that parameters such as age, sex, location and social behavior has no effect on SIR model. The SIRD model also does not consider exposed, super-spreader, and asymptomatic populations. We are presently developing new models to extend and compare with SIRD model, having found insight from the present work. This is particularly not correct in pandemics such as COVID-19 because it was observed that social behavior had great impact for controlling and retarding COVID-19. SIRD model consist of four set of ordinary differential equations (ODE) for susceptible population (S), infected cases (I), and recovered cases (R), and deceased cases (D) [15]. SIRD model is given by [16]:

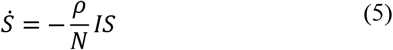

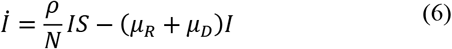

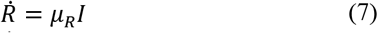

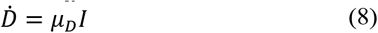

In equations (5-8), there are 4-set of ODE equations based on susceptible population (S), infected population (I), recovered population (R), deceased population (D). Dot-products are time derivatives such as 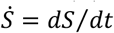.The transmission rate (*ρ*) represent growth rate of infected disease. The recovery rate (*μ*_*R*_) shows the growth rate of recovered population while the death rate (*μ*_*D*_) dictates growth rate of deceased population. The removing rate (*μ* = *μ*_*R*_ + *μ*_*D*_) represents the rate of population removed from susceptible population. The re-production parameter (*R*_0_ = *ρ*/*μ* > 1) is defined to show outbreak of an endemic. Also, *R*_0_ expresses number of contacts from an infected person to susceptible people before complete treatment.

Initial total population (*N*) is assumed a constant number throughout epidemy and is given by:

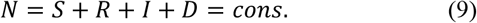

Total infected cases (*TI*) should not be mistaken with active infected population (I), which is described as follows:

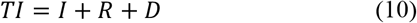

To solve 4-set of ODE equations (5-8), a set of initial conditions are needed as follows (e.g. for total susceptible population of *N*=400,000):

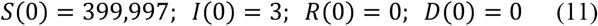

The 4-set of ODE equations (5-8) are solved using MATLAB for time intervals of one day. The set of ODE equations (5-8) are solved using initial conditions (11) in MATLAB for total time duration of contagious disease.

In Wave I, SIRD model 4-set of ODE equations requires initial population of COVID-19 in Iran on February 20, 2020 (the first day of infection) as follows:

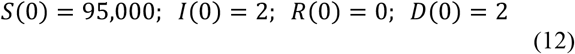

The initial susceptible population, *S*(0), was assumed, infectious, *I*(0), recovered, *R*(0), and dead, *D*(0) were adopted from COVID-19 data of Iran on February 20, 2020 [3].

We consider 5 May 2020 as the start of the Wave II when the infected cases raise to 1,223 on the second wave of COVID-19 in Iran. In Wave II, SIRD model 4-set of ODE equations are solved using initial condition of COVID-19 in Iran as follows:

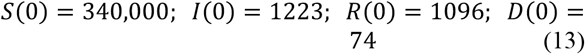

### C. Objective functions

Our viewpoint in this work is different. We used MATLAB optimization to minimize to zero value for the following coefficients (objective function) while comparing model values with COVID-19 clinical data and searching for the optimized best model parameters.

### D. Pearson correlation

Pearson correlation (*R*) is a statistical measure of goodness of fit between M number of model function values (*y*) and COVID-19 data (*z*) as follows [17]:

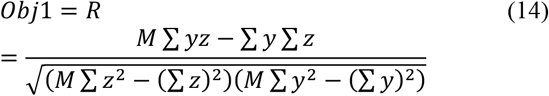

Better fits provide Pearson (*R*) values close to one.

### E. The regression coefficient

The regression coefficient (*R*^*2*^) is another statistical measure for goodness of fit between M number of projected values (*y*) and COVID-19 data (*z*), although we used it as follows [18-19]:

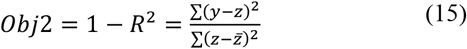

In equation (15), 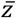 is the average of COVID-19 data and better fits provide the regression coefficient (*R*^*2*^) values close to unity and *Obj2* to zero value

### F. Optimization in MATLAB

To fit model parameters based on available COVID-19 data, the optimization toolbox in MATLAB is used. Fminsearch routine in MATLAB [20] uses a nonlinear programming solver to search for the minimum of an objective function. Routine 1 shows an optimization program in MATLAB to return optimized parameters of a fitting model by minimizing error.

~~~
function error = opt(x,y_covid)
par1=x(1);
par2=x(2);
…
  % Integrate system
  [t,y] =
ode45(@(t,y)YourFunction(t,y,par1,par2,par3,par4),tspan,y0)
…
  % Calculate error
  error = rms(y-y_covid)
end
%% Run optimizer
…
f = @(x)opt(x,y_covid);
[par,fval] = fminsearch(f,x0)
~~~

Routine 1: MATLAB routine to optimize model parameters against COVID-19 data.

As shown in Routine 1, the unknown model parameters par1, par2, … are optimized by minimizing an error estimator to minimize differences between model parameter (y) value against the COVID-19 clinical value (y_covid).

## III. Results

Iran population reported 81.8 million in 2020 [3]. Selecting initial susceptible population S(0) and depends on parameters such as social behavior and government actions to control. This is particularly difficult when some patterns such as double peaks of infections observed. In this study, we divided COVID-19 data of Iran into 2 phases. SIRD model was applied for the first and second expected peaks to assess dynamics of COVID-19 in Iran. Trends of COVID-19 in Iran is presented. The daily and cumulative data of COVID-19 outbreak in Iran are obtained from European Centre for Disease Prevention and Control, COVID-19 situation update worldwide section [21] and are shown in Fig. 1. From daily data, it is observed that Iran has passed the first daily peak after 45 days (April 5, 2020) and reached the minimum after 74 days (May 4, 2020) and then the daily infections has increased towards a second peak after 106 days on June 5, 2020. From cumulative data in Fig. 1, Wave I of COVID-19 in Iran is identified from start of the outbreak on February 20, 2020 to May 4, 2020 with the peak of infectious on April 5, 2020. Wave II is identified from May 5, 2020 to present with the second peak of infectious on June 21, 2020. The daily and cumulative data of COVID-19 outbreak in Iran are shown in Fig. 1.

Next, we applied several mathematical models described in the methodology section for studying Wave I & II of COVID-19 in Iran.

**Figure 1.**
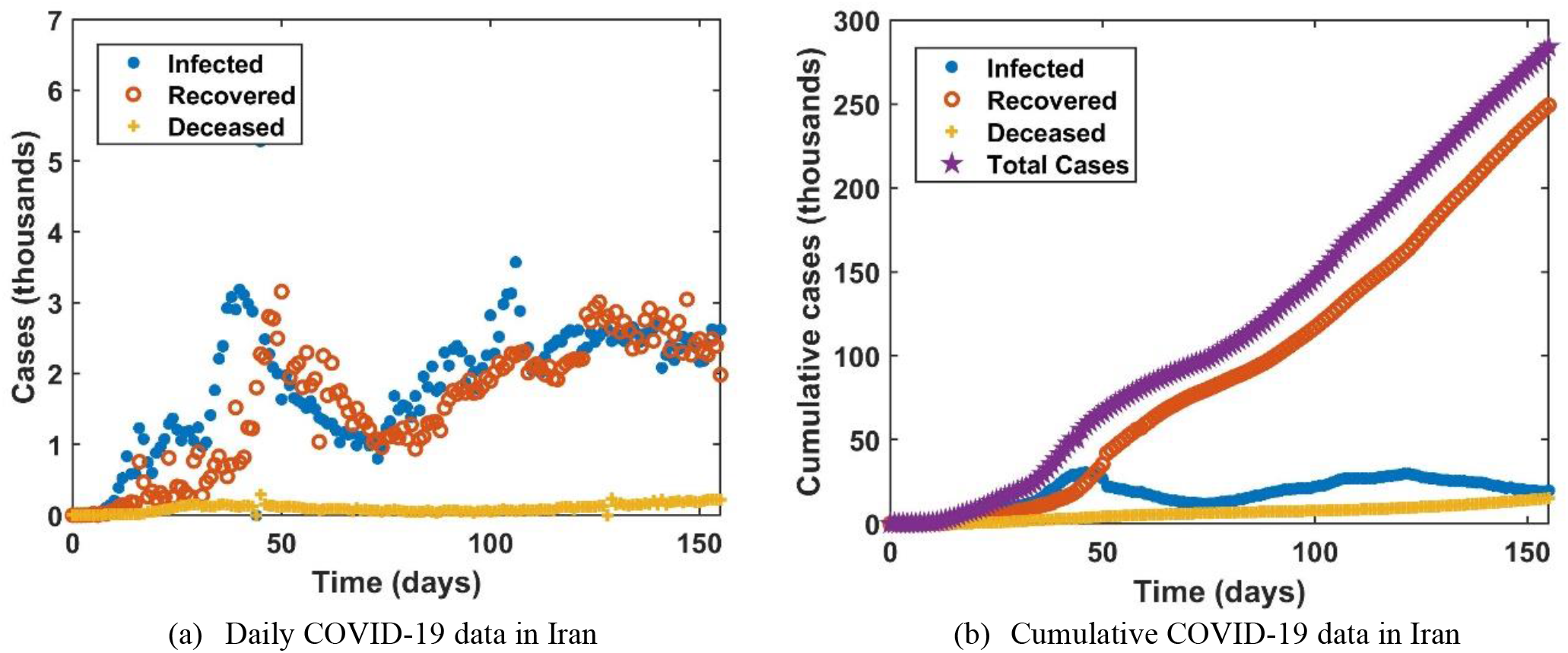
Daily (a) and cumulative (b) data of COVID-19 pandemic in Iran (24 July 2020).

### A. Wave I of COVID-19 in Iran

Wave I of COVID-19 in Iran starts from February 20, 2020 to May 4, 2020. Table 1 summarizes optimum parameters for all models described in methodology section. Accuracy of fitting COVI-19 data with these models are shown by R and R2 values in Table 1.

**Table 1:** Model parameters fitting Wave I of COVID-19 in Iran (24 July 2020).

| Richards Model | $a$ | $b$ | $c$ | $d$ | R | R<sup>2</sup> | |
| --- | --- | --- | --- | --- | --- | --- | --- |
| Total Infected | 102667.56 | 2.8933 | 0.08311 | 0.5519 | 0.9994 | 0.9969 |  |
| Recovered | 92599.73 | 4.0162 | 0.08629 | 0.5944 | 0.9989 | 0.9939 |  |
| Deceased | 6878.31 | 0.005444 | 0.06104 | 0.09095 | 0.9997 | 0.9974 |  |
| Gompertz Model | $a$ | $b$ | $c$ | | R | R<sup>2</sup> | |
| Total Infected | 113870.85 | 2.1940 | 0.05518 |  | 0.9989 | 0.9960 |  |
| Recovered | 111345.09 | 2.7657 | 0.05320 |  | 0.9970 | 0.9914 |  |
| Deceased | 7021.99 | 2.2014 | 0.05660 |  | 0.9997 | 0.9976 |  |
| Logistic Model | $a$ | $b$ | $c$ | | R | R<sup>2</sup> | |
| Total Infected | 98087.84 | 97.4394 | 0.1059 |  | 0.9992 | 0.9965 |  |
| Recovered | 85400.81 | 459.1540 | 0.1162 |  | 0.9988 | 0.9951 |  |
| Deceased | 6166.76 | 86.6776 | 0.1041 |  | 0.9978 | 0.9938 |  |
| Ratkowsky Model | $a$ | $b$ | $c$ | | R | R<sup>2</sup> | |
| Total Infected | 98087.84 | 4.5792 | 0.1059 |  | 0.9992 | 0.9965 |  |
| Recovered | 85400.81 | 6.1294 | 0.1162 |  | 0.9988 | 0.9951 |  |
| Deceased | 6167.99 | 4.4601 | 0.1041 |  | 0.9978 | 0.9938 |  |
| SIRD Model | Transmission rate ($\rho$) | Recovery rate ($\mu_R$) | Death rate ($\mu_D$) | Re-production number ($R_0$) | R<sup>2</sup>(I) | R<sup>2</sup>(R) | R<sup>2</sup>(D) |
|  | 0.36513 | 0.09486 | 0.00765 | 3.56 | 0.8029 | 0.9577 | 0.8854 |

#### Richards Model

Richards model is used to fit total infected (TI), recovered (R), deceased (D) data from Wave I of COVID-19 in Iran. Richards model optimized parameters are listed in Table 1. Fgure 2a shows reslts of Richards model for total ifected cases, recovered, and deceased cases in Wave I of COVID-19 in Iran. As seen in Fig. 2a, it is estimated that the total infected cases will reach to 102,570, recovered cases to 81,587 and fatality to 7,018 by the end of Wave I. On May 4, 2020, these values are 97,424 for total infected cases, 79,379 recovered cases, and 6,203 fatality. Richards model have slightly overestimated total infected cases and recovered yet slightly underestimated fatality in Wave I.

**Figure 2:**
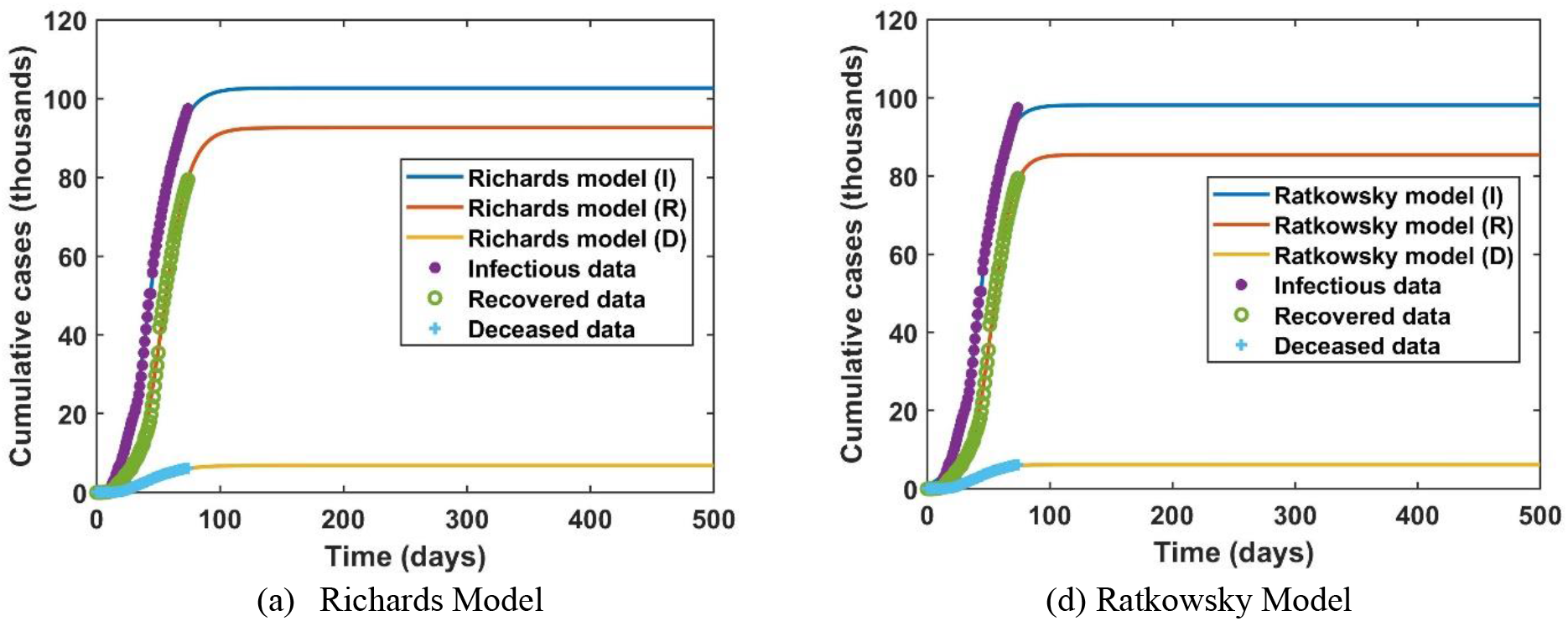
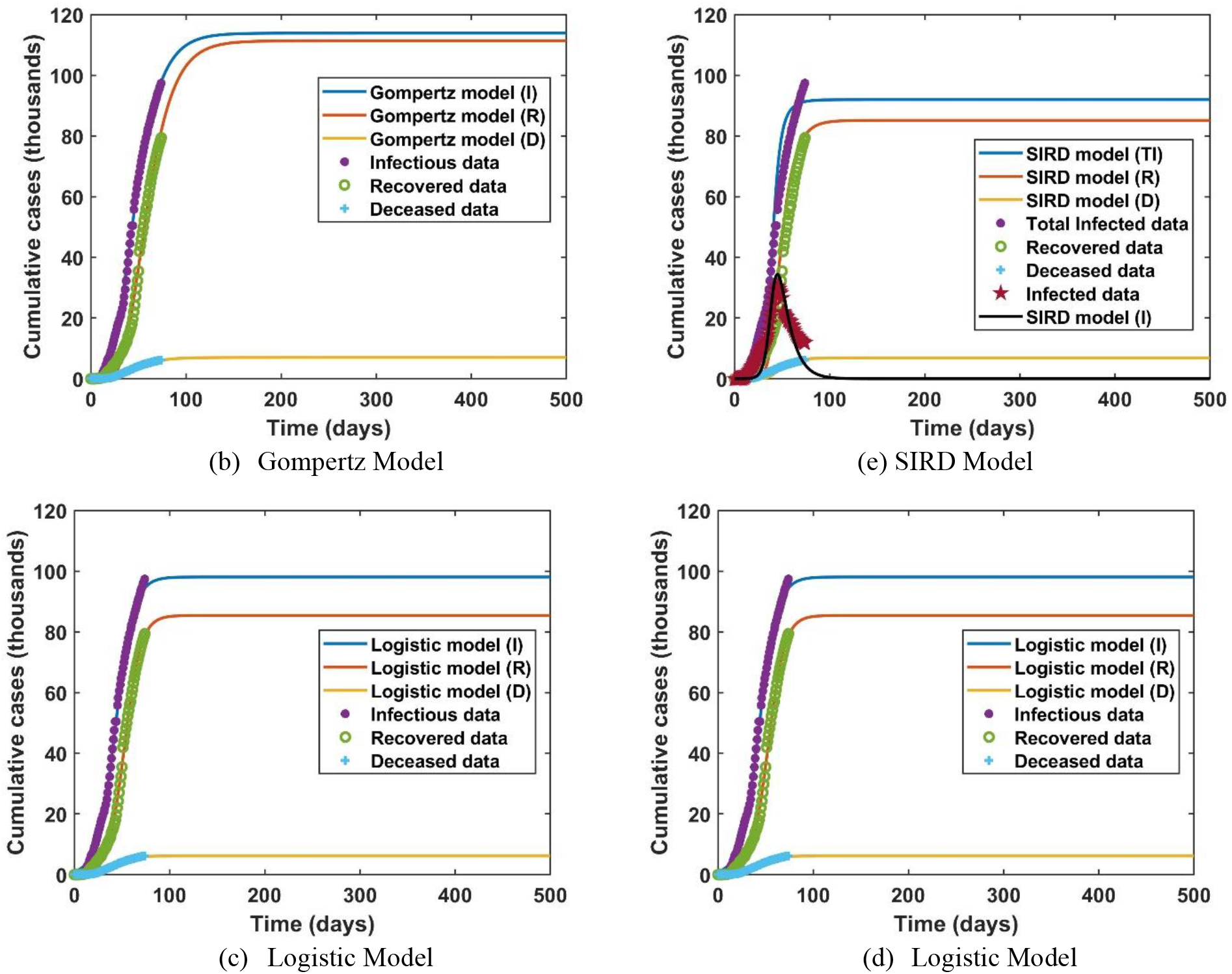
Models projecting trends for Wave I of COVID-19 in Iran (24 July 2020).

#### Gompertz Model

Gompertz model is used here for the Wave I of COVID-19 in Iran with optimized parameters as listed in Table 1. Fgure 2b illustrates results of Gompertz model for estimating Wave I of COVID-19 in Iran. From Fig. 2b, it is observed that total population of infected to reach 113,871 cases, population of recovered to 111,345 cases, and deceased to 7,022 cases by the end of Wave I pandemic in Iran. Comparing these values by actual data on May 4, 2020, i.e. total infected cases of 97,424, recovered cases of 79,379, and deceased cases of 6,203, it is observed that Gompertz model has overeestimated all the populations.

#### Logistic Model

Logistic model is examined here to study Wave I of COVID-19 in Iran. Best fitted values to parameters of Logistic model is listed in Table 1 along with accuracy measures of R and R^2^. Figure 2c illustrates results of Logistic model which indicates total infected cases may reach to 98,088, recovered cases to 85,400, and deceased cases to 6,168 by the end of Wave I pandemic in Iran. Actual data on May 4, 2020 suggest that total infected cases were 97,424, recovered cases 79,379, and deceased cases 6,203. Logistic model has very closely estimated total infected and deceased cases, but recovered cases are overestimated.

#### Ratkowsky Model

Ratkowsky model results are presented for Wave I of COVID-19 pandemic in Iran in Table 1 and Fig. 2d for infected, recovered, deceased cases. As seen in Fig. 2d from February 20, 2020 onwards, Ratkowsky model suggests 98,088 infected cases, 85,400 recovered cases, and 6,168 deceased cases by end of Wave I. Ratkowsky model compares well with actual data on May 4, 2020 consist of 97,424 infected, 79,379 recovered, and 6,203 deceased cases.

#### SIRD Model

Table 1 shows fit parameters of SIRD model to the data. As observed in Table 1, the accuracy of SIRD model to fit COVID-19 data in Iran is acceptable. It is also observed that the reproduction number is relatively high (R0=3.56) in the Wave I of COVID-19 in Iran.

Figure 2e shows the projected results of SIRD model for Wave I of COVID-19 in Iran. As shown in the figure 2e, the peak day of active infectious cases obtained by SIRD model is on April 6, 2020 with 34,447 cases nearly exact match with the actual peak day of infectious which was April 7, 2020 with 30,387 active infected cases. Total recovered cases of 85,117 and total deceased cases of 6,865 can be compared with actual data on May 4, 2020; i.e. 79,379 recovered cases and 6,203 deceased cases. SIRD model is the only model among studied models that can simultaneously fit all 4-set of COVID-19 populations. In addition, SIRD model is the only model to fit peak day of infectious. Although SIRD model has slightly overestimated all exposed populations in Wave I of COVID-19 in Iran.

### B. Wave II of COVID-19 in Iran

We consider May 5, 2020 as the beginning of the Wave II when the infected cases start rising from 1,223 on the second wave of COVID-19 in Iran. Table 2 illustrates model parameters for Wave II optimized to provide accuracies as enlisted by R and R^2^ values.

**Table 2:** Model parameters estimating Wave II of COVID-19 in Iran (24 July 2020).

| Richards Model | *a* | *b* | *c* | *d* | R | R<sup>2</sup> |  |
| --- | --- | --- | --- | --- | --- | --- | --- |
| Total Infected | 245237.13 | -1.2040 | 0.03208 | 0.07686 | 0.9992 | 0.9964 |  |
| Recovered | 255016.26 | -0.3181 | 0.03041 | 0.1487 | 0.9984 | 0.9970 |  |
| Deceased | 51399.22 | -0.001340 | 0.01563 | 0.1428 | 0.9992 | 0.9964 |  |
| Gompertz Model | *a* | *b* | *c* |  | R | R<sup>2</sup> |  |
| Total Infected | 251469.95 | 1.2501 | 0.02976 |  | 0.9985 | 0.9967 |  |
| Recovered | 282636.64 | 1.3555 | 0.02525 |  | 0.9973 | 0.9973 |  |
| Deceased | 65018.52 | 1.6443 | 0.01185 |  | 0.9994 | 0.9966 |  |
| Logistic Model | *a* | *b* | *c* |  | R | R<sup>2</sup> |  |
| Total Infected | 200911.73 | 14.7874 | 0.05958 |  | 0.9972 | 0.9922 |  |
| Recovered | 198723.02 | 19.5511 | 0.05731 |  | 0.9982 | 0.9945 |  |
| Deceased | 17319.55 | 35.3241 | 0.04485 |  | 0.9973 | 0.9959 |  |
| Ratkowsky Model | *a* | *b* | *c* |  | R | R<sup>2</sup> |  |
| Total Infected | 200911.73 | 2.6938 | 0.05958 |  | 0.9982 | 0.9922 |  |
| Recovered | 198723.02 | 2.9730 | 0.05731 |  | 0.9983 | 0.9945 |  |
| Deceased | 17319.55 | 3.5646 | 0.04485 |  | 0.9972 | 0.9959 |  |
| SIRD Model | Transmission rate (*ρ*) | Recovery rate (*μ<sub>R</sub>*) | Death rate (*μ<sub>D</sub>*) | Re-production number (*R*<sub>0</sub>) | R<sup>2</sup>(I) | R<sup>2</sup>(R) | R<sup>2</sup>(D) |
|  | 0.30339 | 0.20026 | 0.00846 | 1.45 | 0.87631 | 0.95694 | 0.94292 |

#### Richards Model

In Wave 2 of COVID-19 in Iran, Richards model results are presented in Fig. 3a. As shown in Fig. 3a, Richards model estimates maximum 245,237 infected cases, 255,016 recovered cases, and 51,399 deceased cases by the end of Wave II of COVID-19 pandemic in Iran. As seen in Fig. 3, Richards and Gampertz models overestimated these populations compared with other models.

**Figure 3:**
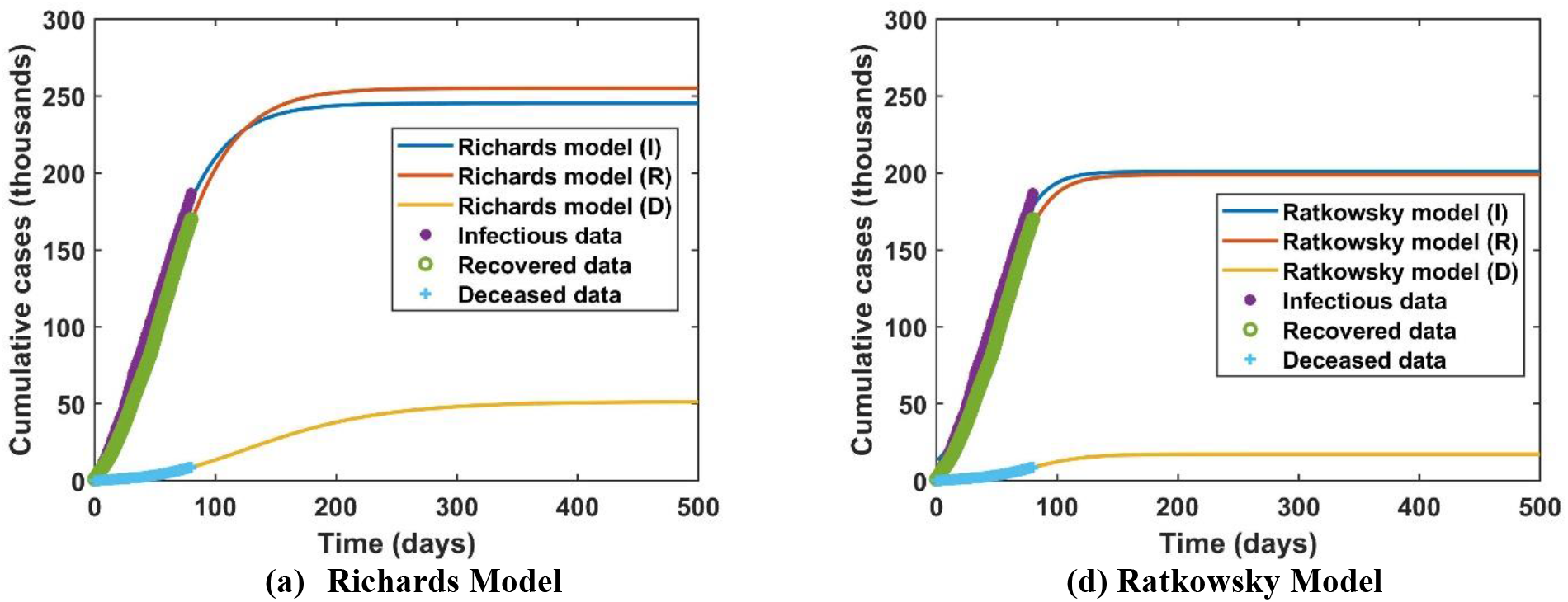
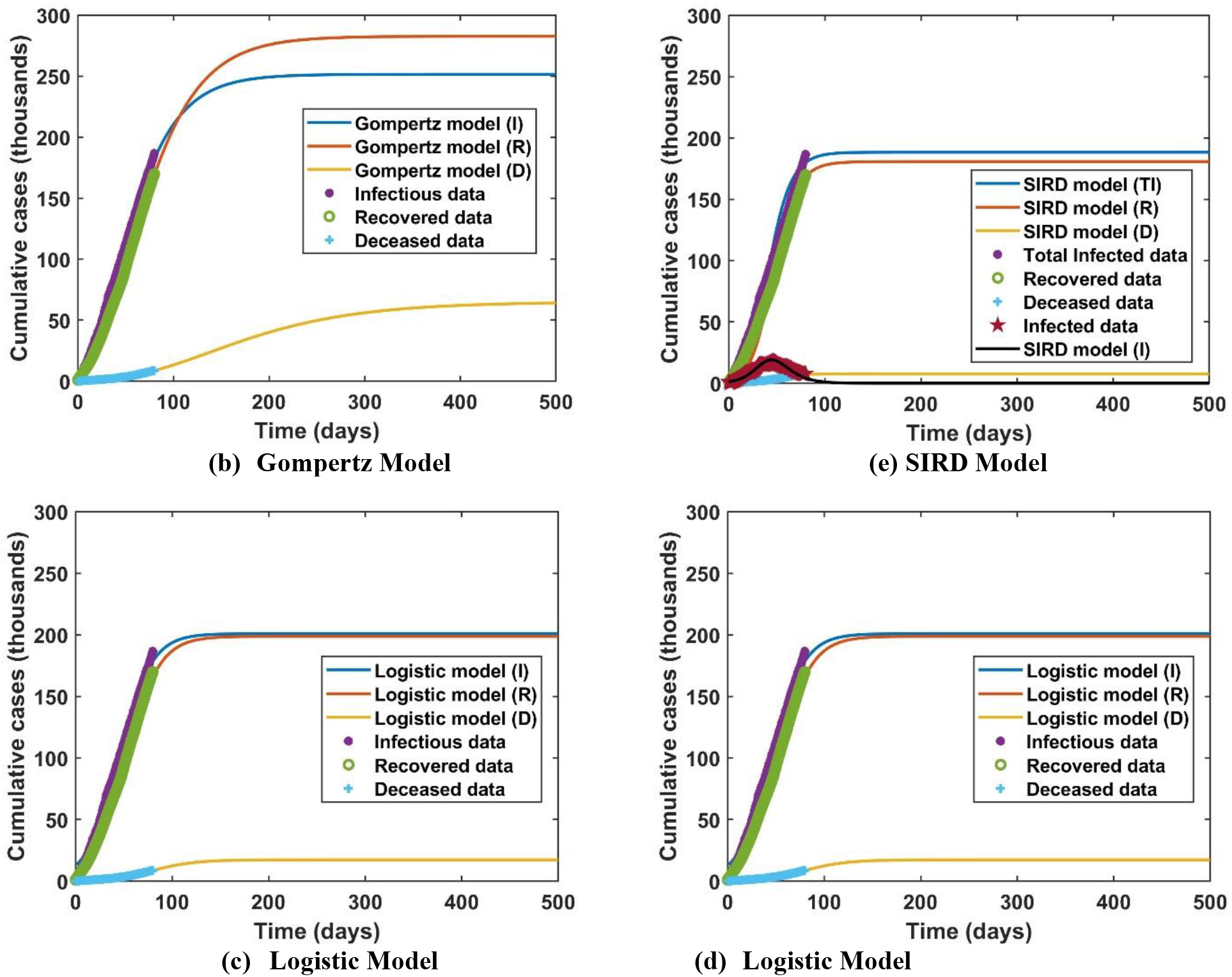
Models estimating trends for Wave II of COVID-19 in Iran (24 July 2020).

#### Gompertz Model

Wave II results of Gompertz model for in Iran is shown Table 2 and Fig. 3b. Fgure 3b illustrates that total population of infected to reach 251,470 cases, population of recovered to 282,636 cases, and population of deceased to 65,018 cases by the end of Wave II. Gompertz model estimations are very high compared with all other models studied here. It is unlikely that Gompertz results on affected populations will be realistic.

#### Logistic Model

Logistic model results for Wave II of COVID-19 in Iran is shown in Table 2 and Fig. 3c. It can be seen from Fig. 3c and Table 2 that Logistic model projects 200,911 infected cases, 198,723 recovered cases, and 17,320 deceased cases for Wave II. Results of Logistic model is closer to Ratkowsky and SIRD models.

#### Ratkowsky Model

Ratkowsky results for Wave II are shown in Table 2 and Fig. 3d for COVID-19 in Iran. Figure 3d shows results from May 5, 2020 to present projecting 200,911 infected cases, 198,723 recovered cases, and 6,168 deceased cases by end of Wave II. Ratkowsky model results compare well with Logistic and SIRD models.

#### SIRD Model

Table 2 illustrates fit parameters of SIRD model to the data. As observed in Table 2, the accuracy of SIRD model to fit COVID-19 data in Iran is very good. In Wave II, the reproduction number (*R*_0_) of 1.45 is obtained. Compare with reproduction in Wave I, it is interesting to see that the second wave of COVID-19 in Wave II is weakened and transmission rate has

## IV. conclutions

It In this work, Richards, Gompertz, Logistic, Ratkowsky and SIRD models are applied to estimate dynamics of COVID-19 in Iran. COVID-19 data from 20 February to 31 May 2020 are used to assess projectability of the studied models on trends of COVID-19 in Iran. The aim is to have a good comparison of widely used models to provide us with even rough projection of future trends of COVID-19 in Iran. This may assist public health organization or policy makers for NPI decisions. There is currently no analytical method to project more waves of COVID-19 beyond Wave II. The outcome of this work may be summarized as follows:

- In Wave I: Richards and Gompertz model overestimated whilst Logistic, Ratkowsky, and SIRD models provided closer projection for COVID-19 data in Iran. WaveSIRD model has projected with nearly exact match the first peak of infection on April 6, 2020 with 34,447 cases (Actual peak day was April 7, 2020 with 30,387 active infected cases).The re-production number (R0) of COVID-19 is calculated 3.95 in Wave I indicating highly transmittable COVID-19 in Iran.
- In Wave II: Richards and Gompertz model very highly overestimated trends whilst Logistic, Ratkowsky, and SIRD models provided close projection to each other.WaveSIRD model provides near exact match for the peak day of infectious; i.e. June 20, 2020 with 19,088 active infected cases compared with actual peak day on June 21, 2020 with 17,644 active infected cases. The re-production number (R0) for Wave II of COVID-19 in Iran is estimated 1.45 indicating that COVID-19 transmission rate was slowed down due to current preventative measure.From results of Wave II, it is observed that the second wave of COVID-19 may affect between 180,000 to 250,000 population and death tool may reach between 6,000 to 65,000 cases depend on different model discussed here.

From results of Wave I COVID-19 in Iran, it is observed that Logistic, Ratkowsky, and SIRD models are better fitting models. Even rough projection of future trends of the selected models for Wave II of COVID-19 may assist in NPI disease management by Iranian government and/or health organization in Iran. There are currently no analytical models available to predict the number of waves and forecast accurately and slowed down. Figure 3e shows the results of SIRD model for Wave II of COVID-19 in Iran. From Fig. 3e, the peak day of infectious is fitted on June 20, 2020 with 19,088 active infected cases whilst actual peak day was June 21, 2020 with 17,644 active infected cases. By November 18, 2020 in the Wave II of COVID-19 in Iran, it is expected that total recovered cases reach to 180,700 whilst total deceased cases reach to 7,655.

completely trends of COVID-19. Richards, Gompertz, Logistic and Ratkowsky are models for only modelling one population. SIRD model is capable of modelling four populations although the model is based on many simplified assumptions that disregard sex, age, location, and behavior of individuals.

## Data Availability

Data is public

https://www.worldometers.info/coronavirus/

